# COVID-19 and Black Fungus: Analysis of the Public Perceptions through Machine Learning

**DOI:** 10.1101/2021.07.08.21260188

**Authors:** Muhammad Nazrul Islam, Nafiz Imtiaz Khan, Tahasin Mahmud

## Abstract

While COVID-19 is ravaging the lives of millions of people across the globe, a second pandemic ‘black fungus’ has surfaced robbing people of their lives especially people who are recovering from coronavirus. Again, the public perceptions regarding such pandemics can be investigated through sentiment analysis of social media data. Thus the objective of this study is to analyze public perceptions through sentiment analysis regarding black fungus during the time of the COVID-19 pandemic. To attain the objective, first, a Support Vector Machine model, with an average AUC of 82.75%, was developed to classify user sentiments in terms of anger, fear, joy, and sad. Next, this Support Vector Machine is used to supervise the class labels of the public tweets (n = 6477) related to COVID-19 and black fungus. As outcome, this study found that public perceptions belong to sad (n = 2370, 36.59 %), followed by joy (n = 2095, 32.34%), fear (n = 1914, 29.55 %) and anger (n = 98, 1.51%) towards black fungus during COVID-19 pandemic. This study also investigated public perceptions of some critical concerns (e.g., education, lockdown, hospital, oxygen, quarantine, and vaccine) and it was found that public perceptions of these issues varied. For example, for the most part, people exhibited fear in social media about education, hospital, vaccine while some people expressed joy about education, hospital, vaccine, and oxygen.

## 1 Introduction

COVID-19 is an infectious disease caused by “Severe Acute Respiratory Syndrome coronavirus 2 (SARS-CoV-2)” which is broadly termed coronavirus [1]. Coronavirus disease has created a global pandemic situation as the death toll continues to rise worldwide. Amidst the crisis of coronavirus, a new epidemic called ‘black fungus’ [2, 3] is spreading fear in people. Black fungus, formally known as mucormycosis, a potentially deadly fungal infection caused by a group of molds called micromycetes. It is more likely to affect people having diabetes, cancer, HIV or aids, and organ transplant that means having compromised immune systems [4]. Cases of mucormycosis have been found in patients who are recovering from coronavirus [5, 6]. As coronavirus leaves its patients’ immune systems in a weakened situation, they are more susceptible to mucormycosis. However, this rare infectious disease is spiraling out of control in India. As of 21 June 2021, 31216 cases of infection and 2109 deaths due to black fungus have been reported [5]. While almost 71% of the global cases of mucormycosis have been reported in India [7].

Due to the COVID-19 outbreak, around half the population of the world was under complete or partial lockdown, which is still ongoing in some countries. To control this outbreak social distancing, staying at home, quarantine is considered the most effective. Thus, social media and social networking sites became very fundamental for expressing opinions and emotions. COVID-19 has altered the way people use the internet since more individuals are logging on to various social media sites. It is possible to comprehend people’s mental states by analyzing their views and opinions, comments, and posts on various platforms. After the surge of coronavirus, some studies have been done focusing on sentiment analysis with Twitter data [8, 9]. During the first phase of the COVID pandemic, false and misinformation were spreading like wildfire. This gave birth to different physiological and mental issues for social media users [10]. The impact of black fungus may affect people in the same way. Moreover, people’s perceptions of black fungus can be explored with the sentiment analysis of social media data.

Machine learning (ML) algorithms learn the hidden pattern of the data and can predict the class labels of unknown samples. Thus, ML is widely being used in the field of health informatics [11] [12] [13] [14], forecasting pandemic [15], predicting shear strength [16], etc. Similarly, ML is widely being used in the field of sentiment analysis for predicting public sentiment [17] [18]. Nonetheless, It can be noticed that a significant number of studies have been conducted focusing on sentiment analysis due to the COVID-19 pandemic in specific as well as cross-country scenarios [19] [10]. However, to the best of our knowledge, the views and feelings of social media users towards black fungus were not revealed yet. Therefore, this article aimed to explore the public views in terms of joy, fear, anger, and sad towards black fungus during the COVID-19 pandemic through sentiment analysis of Twitter data.

## 2 Method

The study was carried out following the phases discussed in the following subsections.

### 2.1 Data acquisition and prepossessing the Twitter data

A total of 8308 tweets were collected through searching Twitter using several keywords for instance ‘COVID-19’, ‘coronavirus’, ‘covid pandemic’, ‘micromycetes’ ‘black fungus’, and ‘COVID delta variant’ in its text. The timestamp of these tweets varies from May 2021 to June 24 since the first black fungus infection case turned up during the COVID-19 pandemic in early May [20]. Then the collected tweets gone through a series of pre-processing steps that subsume: (a) conversion of tweets to lower case character; (b) removal of Username and URLs, punctuation, links, and tabs, white spaces at the start and end of tweets, stop words, (c) expanding of contractions, (d) removal of duplicate tweets. After the pre-processing steps, the reduced dataset contained 6477 tweets.

### 2.2 Developing the machine learning model

Since, the objective of this research was to analyze the public sentiments in terms of joy, fear, anger, and sad [21], a publicly available dataset ^1^ on Twitter that contains Indian sentiment regarding COVID-19, coronavirus, and lockdown, and labeled in terms of joy, fear, anger, and sad was considered for developing the ML model. The collected labeled dataset has also gone through the same set of preprocessing steps as stated in section 2.1. Universal Sentence Encoder (USE) was used for encoding the sentences of the tweets into machine-understandable embedding vectors. A random train test split of 80-20 was done, where 80% data was considered as the training dataset, while 20% data was considered as the test dataset. Next, for Support Vector Machine (SVM) being widely used [22] [23] [24] as well as having better performance [25] [26] for sentiment analysis-related tasks, an SVM model, which is a supervised ML method that can be used to solve classification and regression problems [27], was developed based on the training data to classify user sentiments in the pre-defined class labels. The evaluation of the SVM model is briefly discussed in the Result section.

## 3 Result

### 3.1 Analyzing the performance of the SVM model

The performance of the developed ML model is analyzed based on precision, recall, and f1 score. The precision, recall, and f1 score for the training dataset were 93.0%, 93.1%, and 93.0% respectively and for the test dataset, the performance measures were 73.8% precision, 74.2% recall, and 73.8% f1-score. The ROC Curves and Confusion matrices are also generated for both the train as well as the test data to further analyze the model. ROC Curves for both the train and test dataset are shown in Figure 1(a) and 1 (b) respectively, while Confusion matrices for the train and test dataset are shown in Figure 2(a) and 2(b) respectively. In Figure 1 and 2, label 0, 1, 2, and 3 represent anger, fear, joy, and sad respectively. It can be seen from Figure 1 (b) that, for the test data, Area Under the Curve (AUC) for class labels 0, 1, 2, and 3 were 80%, 77%, 89%, and 85% respectively (82.75% average AUC), while AUC for class labels 0, 1, 2, and 3 were 95%, 93%, 97% and 97% respectively (95.5% average AUC) for the train dataset (see Figure 1 (a)). However, it can be seen from the right diagonals of figure 2(b) that, 113, 111, 118, and 112 test samples were correctly classified as labels 0, 1, 2, and 3 respectively, while 566, 547, 565, and 622 train samples were correctly classified as 0, 1, 2 and 3 respectively. Therefore, it can be said from the ROC Curves and the confusion matrices that the model has achieved satisfactory performance for both the train as well as the test dataset.

**Figure 1:**
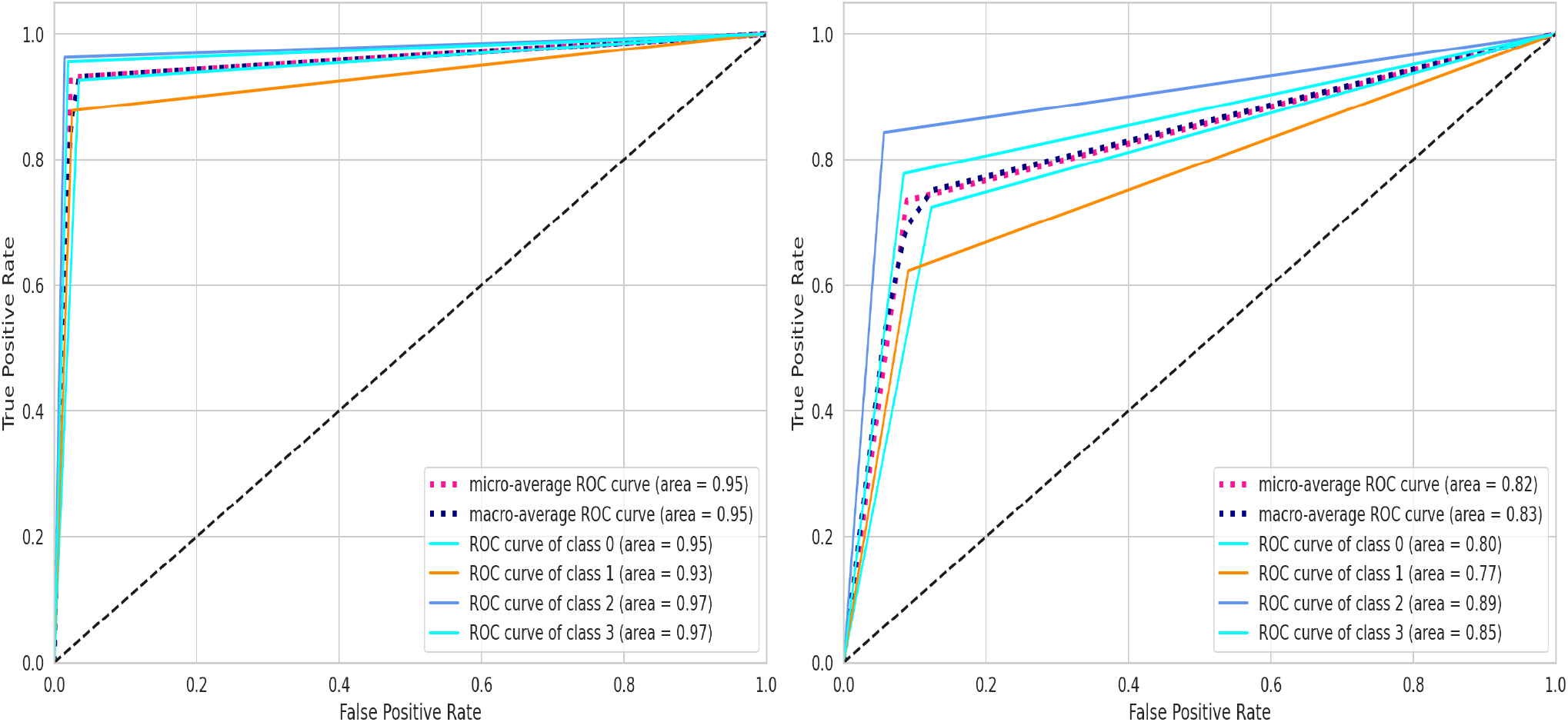
ROC Curves for the developed SVC model: (a) train data, (b) test data

**Figure 2:**
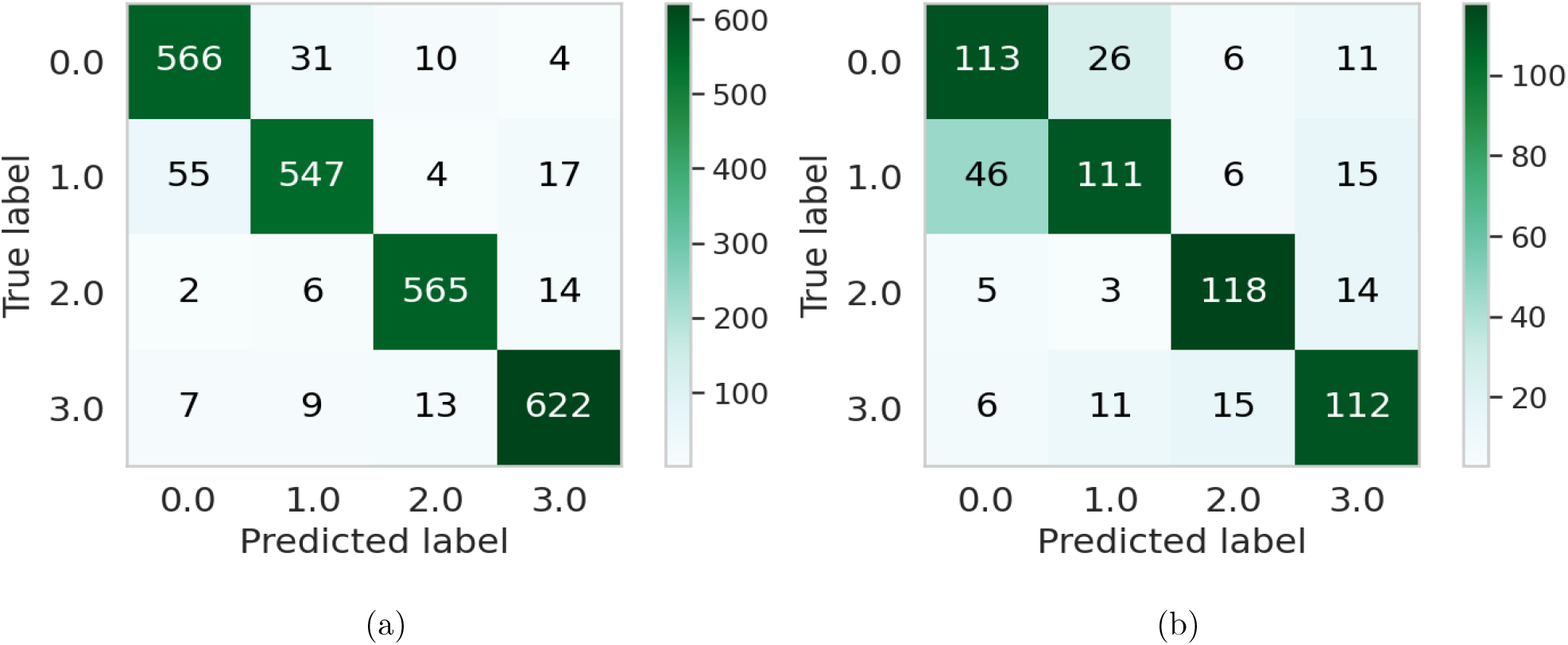
Confusion matrices for the developed SVC model: (a) train data, (b) test data

### 3.2 Analysis of public sentiments

The trained ML model was used to predict the sentiments of the 6477 unlabeled tweets (as stated in section 2.1). Some of the tweets and their model predicted sentiments are shown in Table 1. The model predicted classified data are analyzed for understanding the public perceptions towards black fungus during the COVID-19 outbreak.

**Table 1:** Example tweets regarding COVID-19 and Black Fungus

| Tweet No. | Tweet | Sentiment |
| --- | --- | --- |
| 1 | False claims about black fungus have flooded social media platforms. Many believe that black fungus is caused by wearing masks, or that it is a symptom of the new COVID-19 strain in India. | Anger |
| 2 | The green fungus case has induced a fresh wave of fear across India, which over the year has extensively witnessed cases of black, white and yellow fungus following COVID-19 infections. | Fear |
| 3 | Potentially fatal black fungus cases reported in covid-19 patients in Oman, as infections there surge. | Fear |
| 4 | India has reported the first case of green fungus in a covid-19 recovered patient in Madhya Pradesh. The country has already registered cases of black fungus, white fungus and yellow fungus. | Sad |
| 5 | Dutch kit helps detect Mucormycosis(Black fungus) in 24-48 hrs. instead of 10-15 days of current days, said Dr. K Bhujang Shetty, Chairman NNethralaya It is equivalent to RT-PCR test for COVID-19 & 5 varieties of black fungus can be detected by this kit. | Joy |
| 6 | Oman reports new cases of black fungus in COVID-19 patients following 'epidemic' of the infection in India. | Sad |

The classification of tweets into four sentiments is displayed in Table 2. It can be seen from Table 2 that, 36.59% of people expressed sadness, followed by joy (32.34%), fear (32.34%), and anger (1.51%). Surprisingly, results showed more tweets expressing joy, than fear. However, it was observed that lots of people expressing their positive sentiment after getting vaccines, after getting cured of COVID-19, being optimistic to fight against the pandemic, etc, which could be the reason for finding more joyful tweets than fearful tweets.

**Table 2:** Sentiments labels of the classified Tweets

| Sentiment | No. of Tweets | Percentage |
| --- | --- | --- |
| anger | 98 | 1.51% |
| fear | 1914 | 29.55% |
| joy | 2095 | 32.34% |
| sad | 2370 | 36.59% |

It is apparent that in the majority of the tweets (36.59%) people were expressing sad which is natural since the COVID pandemic has already put mental stress on people of all ages. On top of this, black fungus infections have added an extra layer of mental burden and thus of the tweets were expressing sadness. However, only a few tweets can be found expressing anger.

### 3.3 Analysis of recurring words in word clouds

The word clouds for anger, fear, joy, and sad sentiments are presented in Figure 3. The result showed that fungus, covid, black and white are recurring in each of the sentiments (see Figure 3 (a,b,c,d)). In the case of anger sentiment (see Figure 3 (a)) the most frequent words are fungus, black, covid, government, cancel, white, and virus. This indicates the public’s dissatisfaction with the government initiatives taken to restrain the black fungus infection during the COVID-19 pandemic. Again, the words fungus, black, infection, covid, deadly, disease, new, rise, warn, found, diabetes, infection, suffering were found in expressing fear and sadness (see Figure 3 (b, d)). These words indicate that people are concerned about the fact that COVID-19 and diabetes leave the immune system of patients in a vulnerable state [28], which results in people being at a higher risk of black fungus infection. Cases of black fungus developed in people with COVID-19 with increased blood-sugar levels were high[29]. Also, people think that black fungus is a new deadly infectious disease even though it has been around for centuries [30]. This is because of the recent emergence of black fungus amidst covid pandemic, so the words new, deadly, disease were found in the word clouds of negative sentiments (fear and sad) (see Figure 3 (b, d)). Moreover, the words white and yellow were found in all four of the word clouds since white and yellow fungus are more rarer and lethal fungal diseases than black fungus [31].

**Figure 3:**
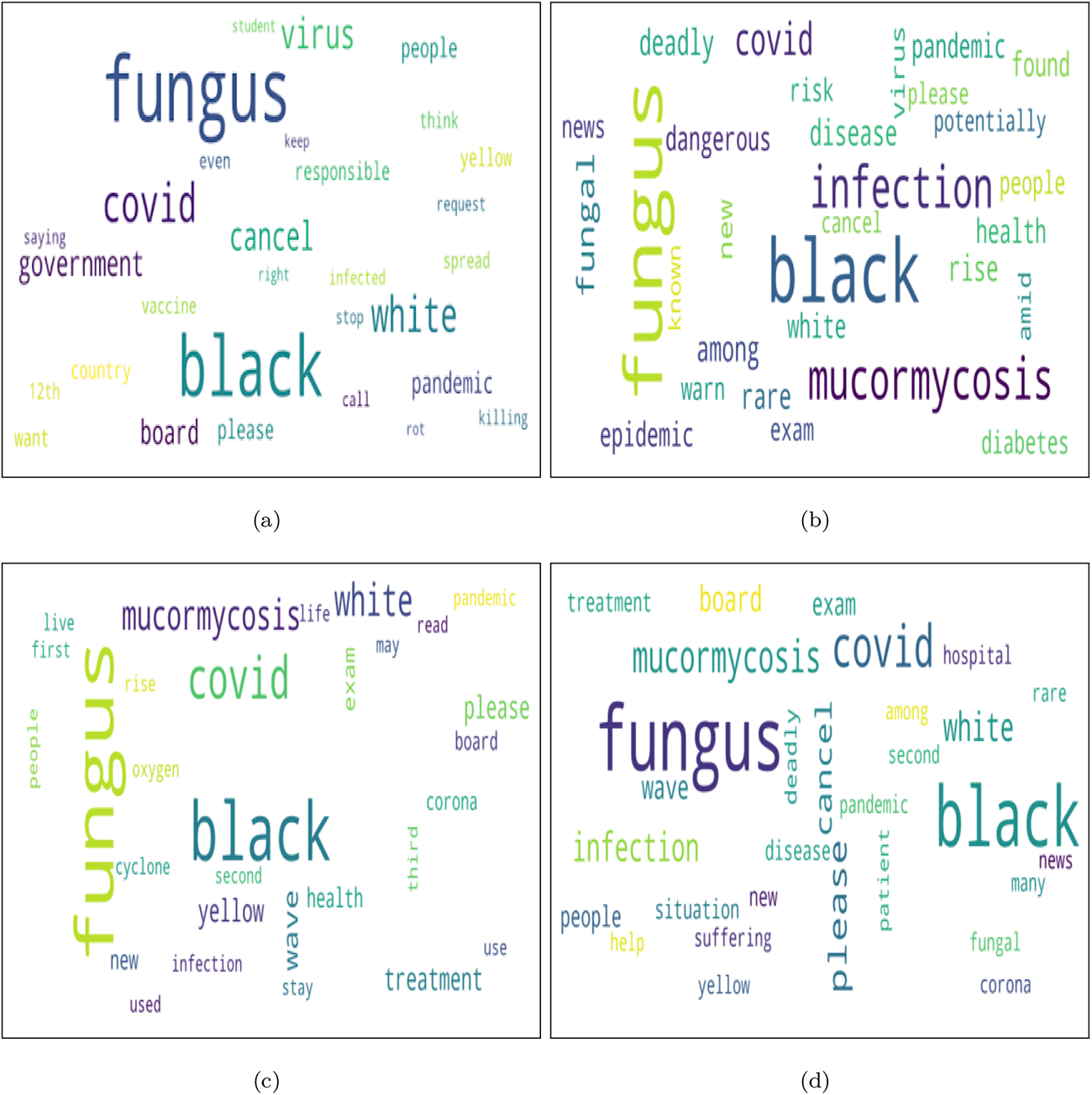
Word clouds generated from sentiments: (a) anger, (b) fear, (c) joy, and (d) sad

### 3.4 Public sentiment regarding specific concerns

To provide a better understanding of people’s perspective on black fungus, few concerns were highlighted based on the related recurring words, and the sentiments (anger, fear, sad, and joy) were analyzed regarding these concerns. Figure 4 depicts the number of tweets to different concerns and people’s sentiment to each concern. Education, vaccination, and hospital-related tweets had more negative sentiments (fear and sadness) than positive sentiments, whereas hospital and vaccine-related tweets have more sentiments expressing sadness than the other three sentiments. This indicated that people’s perceptions of healthcare management to handle black fungus and COVID-19 patients during this pandemic were not satisfactory. During the coronavirus pandemic, hospital management systems in several nations crumbled due to the unrestrained rate of COVID - 19 infection [32]. Many hospitals were even forced to turn away patients in dire need. People expressed fewer positive feelings (joy) about education, hospitals, quarantine, oxygen, and vaccines than negative feelings. Since people’s attention has turned away from lockdown, few tweets on lockdown and quarantine have been detected. This also indicates the ignorance tendency of people to lockdown, COVID-19 restrictions, and prescribed hygiene rules though the coronavirus and black fungus infections rates broke the previous infection records many times.

**Figure 4:**
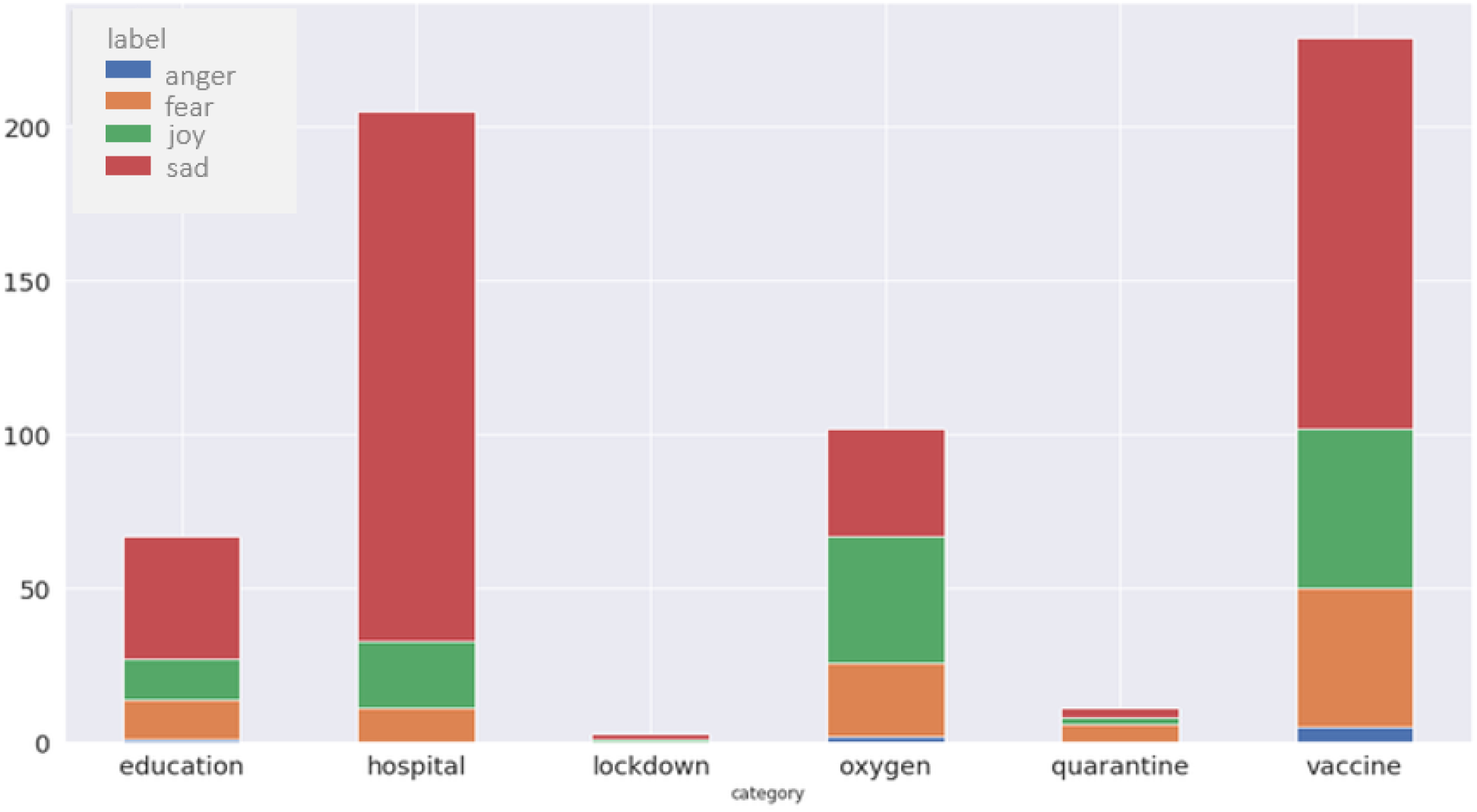
Public sentiment regarding specific concerns

## 4 Conclusion

In this study, data extracted from Twitter was mined to understand the public sentiments towards black fungus during the time of the COVID-19 pandemic. A Support Vector Classifier was used for the classification of tweets into sad, joy, fear, and anger sentiments. The classifier had an average AUC of 82.75% on the test dataset. As outcome, the public perceptions were distributed into four sentiments as follows: sad (n = 2370, 36.59 %), joy (n = 2095, 32.34%), fear (n = 1914, 29.55 %) and anger (n = 98, 1.51%). Then this study also revealed public perceptions on several important concerns, in particular, education, lockdown, hospital, oxygen, quarantine, and vaccine. It was also found that regarding the topics on education, hospital, oxygen, quarantine, and vaccine people had more negative feelings (fear and sadness) than positive feelings while people paid almost no attention regarding lockdown and quarantine. These findings indicate that people are not more likely to stay at home, maintaining social distancing in this pandemic. Thus many countries in this world are found to be using many synonymous words of Lockdown like shutdown, strict lockdown; just to motivate folks to stay at home [33] [34] [35]. However, this study findings can provide a deeper understanding of public perceptions towards black fungus during the COVID-19 epidemic in the world. Moreover, this study can help the government and policymakers to takes important decisions and actions for controlling the black fungus and COVID-19 outbreaks.

## Data Availability

Data used in this article is not made public by the authors

## Footnotes

1 https://www.kaggle.com/surajkum1198/twitterdata

